# Age-dependent acceleration of structural brain aging in medication-free major depressive disorder linked to neuroanatomical phenotype findings from COORDINATE-MDD consortium

**DOI:** 10.64898/2026.03.31.26349338

**Authors:** Bhanu Sharma, Pedro L. Ballester, Luciano Minuzzi, Wenyi Xiao, Mathilde Antoniades, Dhivya Srinivasan, Guray Erus, Jose A. Garcia, Yong Fan, Danilo Arnone, Stephen R. Arnott, Taolin Chen, Ki Sueng Choi, Katharine Dunlop, Cherise Chin Fatt, Rachel D. Woodham, Beata R. Godlewska, Stefanie Hassel, Keith Ho, Andrew M. McIntosh, Kun Qin, Susan Rotzinger, Matthew D. Sacchet, Jonathan Savitz, Haochang Shou, Ashish Singh, Vibe G. Frokjaer, Melanie Ganz, Aleks Stolicyn, Irina Strigo, Duygu Tosun, Dongtao Wei, Ian M. Anderson, W. Edward Craighead, J. F. William Deakin, Boadie W. Dunlop, Rebecca Elliott, Qiyong Gong, Ian H. Gotlib, Catherine J. Harmer, Sidney H. Kennedy, Gitte M. Knudsen, Helen S. Mayberg, Martin P. Paulus, Jiang Qiu, Madhukar H. Trivedi, Heather C. Whalley, Chao-Gan Yan, Allan H. Young, Christos Davatzikos, Cynthia H. Y. Fu, Benicio N. Frey

## Abstract

**Background:** Major depressive disorder (MDD) is associated with altered brain structure and evidence of accelerated brain aging. However, previous studies have been limited by clinical samples with mixed medication status and multiple mood states, modest sample sizes, small percentage of MDD individuals older than 65 years of age, and/or reliance on summary-level data.

**Methods:** Harmonized T1-weighted MRI from MDD (n = 645), all medication-free and in a current depressive episode, and matched healthy controls (n = 645), segmented into 145 regional volumes, from 11 sites in COORDINATE-MDD consortium. Brain age gap (BAG) was estimated using gradient boosting regression with nested cross-validation. Group differences in BAG (and age-corrected BAG [cBAG]) were examined across age strata. Regional contributions were evaluated using Shapley Additive exPlanations.

**Results:** MDD was associated with significantly elevated cBAG compared with healthy controls (mean difference + 2.01 years). Age-stratified analyses showed no differences before mid-30s, with progressively larger gaps thereafter, reaching +6.85 years in MDD aged 55 and older. cBAG differed across neuroanatomical phenotypes associated with differential antidepressant response, cognitive impairment, increased adverse life events, increased self-harm and suicide attempts, and a pro-atherogenic metabolic profile. Key contributing regions included lateral and medial prefrontal regions, middle temporal gyrus, putamen, supplementary motor cortex, central operculum, and cerebellum.

**Conclusions:** Accelerated structural brain aging in MDD is age-dependent and is most pronounced in a neuroanatomical phenotype associated with worse key clinical outcomes. The findings support neuroprogression models of MDD while demonstrating that cBAG is not a uniform feature of MDD and seem to be more strongly expressed in a specifically clinically vulnerable disease phenotype.

## INTRODUCTION

Major depressive disorder (MDD) is a prevalent psychiatric condition affecting over 250 million individuals worldwide^1^. Meta-analytic findings based on over 90 studies (including over 1 million adults) from 30 countries estimate a lifetime prevalence exceeding 10%, reaching nearly 15% in women^2^. Despite its substantial impact, MDD remains a clinically defined syndrome, lacking a clear neurobiological etiology^3^. Diagnostic criteria and core features of MDD include low mood and/or anhedonia, accompanied by disturbances in sleep, energy, cognition and psychomotor functioning, and in some cases, suicidal ideation and behaviors^3,4^. While these symptoms are common to MDD, its clinical presentation varies by age and sex^5^. Beyond the individual burden, MDD imposes considerable societal and economic costs, contributing to increased healthcare utilization^6^ and an estimated $400 billion in annual economic expenditure in the United States alone^7^. With the World Health Organization predicting that MDD will become the leading burden of disease worldwide by 2030^8,9^, advancing neurobiological models of depression is essential for improving diagnosis, treatment, and long-term management.

Neuroimaging studies have identified MDD as a disorder marked by “accelerated aging”^10^, wherein volumetric reductions are observed in one or more anatomical regions. The application of machine learning algorithms to neuroimaging data allows for estimation of an individual’s brain-predicted age based on structural or functional features, such as neuroanatomical regions of interest (ROIs). The difference between brain-predicted age and chronological age is known as the brain age gap (BAG), which provides a quantitative index of deviation from normal aging trajectories, whether brain aging is accelerated^11,12^.

A meta-analysis of 7 studies (with a total MDD sample of 3423 and a control sample of 2874, ranging in individual study samples from 139 to 4801) found that a mean BAG difference of 1.12 years (95% CI: 0.41-1.83)^10^. Importantly, increased BAG in older adults with MDD is associated with poorer cognitive performance (mainly executive functioning and working memory), suggesting that it may be a translational marker ^13^. Converging evidence from non-MDD populations further demonstrate associations between elevated BAG and impairment in multiple cognitive domains measures ^14^ indicating that BAG may be a marker of clinically meaningful brain alterations.

However, BAG studies in MDD have been limited by modest sample sizes^10^, mixed medication status^15,16^, summary-level data^16,17^, and lack of age stratification. While Enhancing NeuroImaging Genetics through Meta-Analysis (ENIGMA) consortium consisted of a large sample (n>6000)^16,17^, it included subjects with a wide range of clinical presentations, limited medical history, mixed medication status, including antidepressant/psychotropic medication usage, and multiple co-morbidities^18^. Further, in using summary-level data, it was recognized by the authors that heterogeneity across scan sites was a potential reason for their relatively small, pooled effect size with BAG ranging from 0.99 to 1.08 years^17^. These confounds need to be addressed to continue to improve our understanding of BAG in MDD. In addition, prior studies in MDD have offered few insights into which ROIs contribute most to BAG predictions, limiting our ability to understand which regions or networks may be disproportionately structurally impacted.

The present study addresses these limitations by leveraging individual-level, harmonized structural MRI data in medication-free individuals with current MDD from the COORDINATE-MDD consortium^19^, which addresses key methodological limitations of prior large-scale neuroimaging efforts. Our objective was to quantify the BAG in MDD relative to healthy controls, determine whether brain aging varies across the lifespan, and identify the individual ROIs that most strongly contribute to brain age deviations. We also investigated BAG in neuroanatomical phenotypes associated with differential antidepressant treatment response and clinical and cognitive profiles^19,20^.

## METHODS

### Participants and data collection

This study is a product of the international consortium COORDINATE-MDD^21^. Participants in this study were required to meet the following inclusion criteria: Diagnostic and Statistical Manual of Mental Disorders or the International Classification of Diseases (ICD) diagnosis of MDD, and in a current medication-free episode of depression of at least moderate severity (or 14+ points on the Hamilton Rating Scale for Depression)^21,22^. Participants were excluded if they had treatment-resistant depression or comorbid psychiatric, medical, or neurological disorders.

### MRI processing

T1-weighted images (MPRAGE or equivalent structural sequences) were acquired from all sites on either 1.5T or 3.0T magnets; multiple MRI vendors were used. Given the hardware and acquisition protocol differences across sites, data harmonization was performed on raw images^23^. This was achieved using a general additive model (GAM) combined with the existing COMBAT method to harmonize data across sites^21^.

Each T1-weighted image was quality checked (for motion, image artifacts, and/or restricted field-of-view) and then processed using a standardized pipeline, which involved magnetic field inhomogeneity correction^24^ and multi-atlas skull-stripping^25^. Further, we segmented images using a multi-atlas approach, namely the Multi-Atlas Region Segmentation Using Ensembles (MUSE), a robust method that allows for non-linear registration to each atlas in the ensemble, and then a label-fusion approach that is used to assign a final label to each voxel in the T1-weighted image based on the aggregation of labels^26^. MUSE then provides a mix of single ROIs (or distinct neuroanatomical regions) or composite ROIs which include multiple, individual anatomical regions. For the purposes of our analyses, only the single ROIs were included in the analysis. All ROIs were normalized to ICV.

### Propensity score matching

Prior to modelling, to reduce confounding, propensity score matching was applied using age (z-scored) and ICV (z-scored) as covariates. Logistic regression was used to estimate propensity scores, and participants were matched 1:1 using k-nearest neighbor matching with a 0.5 SD caliper and no replacement.

Ultimately, this present study included 1290 adults (645 with MDD and 645 healthy controls) across 11 international sites participating in the COORDINATE-MDD consortium. The sites that contributed to the overall sample included: Establishing Moderators and Biosignatures of Antidepressant Response in Clinical Care (EMBARC), n=270; Huaxi MR Research Center at Sichuan University (SCU), n = 250; Laureate Institute for Brain Research (LIBR), n = 195; Stratifying Resilience and Depression Longitudinally (STRADL), n = 135; Canadian Biomarker Integration Network in Depression (CAN-BIND), n = 93; the Manchester Remedi cohort, n = 70; the Oxford cohort, n = 70; Predictors of Remission in Depression to Individual and Combined Treatments (PReDICT), n = 63; the Stanford cohort (SNAP), n = 58; the Manchester Blame cohort, n = 46, and the King’s College London cohort (KCL), n = 40. The sample distribution is reported in **Supplementary Table 2** and the demographics are reported in **Table 3**.

The principal investigators of each of member site also provided anonymized demographic and clinical data to the consortium. Detailed methods regarding recruitment and clinical phenotyping are reported in the consortium’s published protocol^21^. All sites acquired ethical approval prior to study initiation: Manchester (Stockport Research Ethics Committee 07/H1012/76), SNAP (IRB approval 12104), EMBARC (STU 092010–151), Oxford (REC reference 11/SC/0224), LIBR (WCG IRB 1136261 and 1136947), STRADL (NHS Tayside committee 14/SS/0039), PReDICT (Emory IRB # 00024975), KCL (Bromley NHS REC 13/LO/0904), CANBIND (11-0917-A) and SCU (IRB 2020(54)).

### Model parameters

A gradient boosting regression model was implemented using scikit-learn’s GradientBoostingRegressor to predict brain age from ROI volumes. The hyperparameter search space was designed to optimize model performance while preventing overfitting, encompassing eight key parameters: number of estimators (100-500), learning rate (0.001-0.3), maximum tree depth (2-7), minimum samples per split (2-15), minimum samples per leaf (1-6), subsample ratio (0.6-1.0), feature selection method (sqrt, log2, or all features), and loss function (squared_error, absolute_error, huber, or quantile).

To ensure robust model evaluation and prevent data leakage, a nested cross-validation approach was implemented with 5-fold outer and inner loops. The outer loop provided unbiased performance estimates on held-out control data, while the inner loop performed hyperparameter optimization using randomized search with 50 iterations per fold. This dual-loop structure ensured that hyperparameter selection occurred independently within each outer fold, preventing overfitting to the specific data splits used for final evaluation. Model performance was evaluated using MAE.

Further, to enhance model interpretability and identify the most influential brain regions for brain age prediction, we computed SHAP values using the TreeExplainer from the SHAP library (v0.42.0). SHAP values provide a unified framework for explaining individual predictions by quantifying the contribution of each feature to the final model output, ensuring that the sum of all feature contributions equals the difference between the predicted value and the expected value. SHAP values were calculated using out-of-fold predictions to ensure unbiased estimates of feature importance. This approach quantifies the contribution of each ROI to the brain age prediction. The mean absolute SHAP values were used to rank ROIs by their importance, providing insights into which neuroanatomical structures are most predictive of brain age. This interpretability framework enables both global feature importance ranking and individual-level explanations, facilitating the identification of brain regions that may be most sensitive to age-related changes and potentially altered in MDD.

To correct for systematic bias in brain age predictions (e.g., overestimation in younger individuals, underestimation in older), we applied linear correction to predicted age and BAG. First, predicted brain age was regressed on chronological age in the control group to compute residuals: cPRED = PRED – (β₀ + β₁ × age), where β₀ and β₁ are the intercept and slope coefficients from the linear regression of predicted age on chronological age in the control group. The corrected BAG was then defined as: cBAG = cPRED – age. These corrections remove age-related dependency and allow for unbiased estimation of brain age deviation. This approach assumes that the control group represents the normative aging trajectory, and any deviation from this trajectory in clinical groups reflects true brain age acceleration rather than systematic prediction bias. The correction effectively centers the BAG around zero for the control group, providing a reference point for interpreting accelerated or decelerated brain aging in clinical populations.

### Statistical analysis

To examine the relationship between BAG and MDD, we conducted several statistical analyses. All analyses were conducted using Python 3.13.3 with scipy, statsmodels, and scikit-learn libraries, with statistical significance set at p< 0.05 (FDR-corrected where applicable).

Propensity score matching was performed to balance age and ICV between MDD and control groups, ensuring comparable demographic and anatomical characteristics. Balance between groups was assessed using independent t-tests (age, ICV) and chi-square tests (sex, which was not propensity matched as the groups were already comparable with respect to this variable).

For feature-level analysis, we examined the interaction between diagnosis and individual ROIs using Ordinary Least Squares (OLS) regression models: cBAG ∼ diagnosis + ROI + diagnosis:ROI, where ROI represents individual brain region volumes. This approach identifies ROIs that show differential associations with brain age gap between diagnostic groups. To control for multiple comparisons across the large number of brain regions, we applied False Discovery Rate (FDR) correction using the Benjamini-Hochberg procedure (α = 0.05).

Group comparisons were conducted using independent t-tests to compare cBAG between MDD and control groups, with effect sizes calculated using Cohen’s d. Age-stratified analyses were performed by dividing participants into age groups (18-25, 26-35, 36-45, 46-55, 55+ years) and conducting independent t-tests for diagnostic differences within each age stratum, with effect sizes computed for each comparison. Based on our consortia’s prior work, we identified two MDD neuroanatomical phenotypes, or Dimensions, that predict treatment response^50^. The first of these Dimensions was characterized by relatively preserved grey and white matter, while the second showed widespread but subtle volume losses in comparison to controls. As such, we also examined cBAG across these previously identified MDD dimensions.

Site effects were assessed using Intraclass Correlation Coefficient (ICC) and Leave-One-Site-Out (LOSO) cross-validation, in which models were iteratively trained on data from all but one imaging center and tested on the held-out site, to evaluate generalizability across scanners and acquisition protocols. Uncertainty estimation was provided through bootstrap confidence intervals (10,000 resamples) for mean differences and effect sizes to provide robust uncertainty estimates.

## RESULTS

### Normalization and propensity score matching

Structural MRI data on 693 and 669 controls and MDD subjects were imported from a harmonized multi-site dataset. Only individual ROIs volumes were retained for the present analysis, excluding derived or composite regions. All ROI volumes were normalized by intracranial volume (ICV) to account for individual differences. The final dataset comprised 145 ICV-normalized ROI volumes per participant, providing a comprehensive representation of brain structure.

Propensity score matching was performed using logistic regression (caliper width = 0.5) to match MDD subjects with healthy controls based on age and ICV. Following matching, 645 subjects were retained per group. Statistical tests confirmed balance between groups: independent t-tests revealed no significant differences in ICV (t=0.063, p=0.950) or age (t=1.085, p=0.278) between matched controls and MDD subjects. Additionally, a *post-hoc* Chi-square test confirmed no significant difference in sex distribution between groups (χ^2^=1.184, p=0.276).

### Model performance and cross-validation

We used a gradient boosting regression model to predict brain age from ROI volumes. Nested cross-validation (5-fold outer and inner loops) on the control sample yielded consistent performance across all outer folds, with individual fold MAE values ranging from 6.79 to 8.13 years (**Supplementary Table 1**). The mean MAE across all outer folds was 7.379 ± 0.454 years, demonstrating stable model performance with relatively low variance between folds. The best-performing model emerged from outer fold 3, achieving an MAE of 6.788 years. After nested cross-validation provided an unbiased estimate of generalization performance, we retrained a final model on the complete control dataset using the same hyperparameter optimization procedure. This final model was then applied to the MDD test set, achieving an MAE of 6.540 years. The performance on the MDD test set suggests that the model generalizes well to the clinical population.

### Feature importance

Shapley Additive exPlanations (SHAP) analysis was used to quantify the contribution of each feature (or ROI) to brain age prediction, with the top 20 most important features identified. Interaction effects (Group X Feature) were tested with an ordinary least squares (OLS) regression, with several ROIs surviving multiple comparisons correction. Specifically, for prediction of both corrected measures (namely, corrected predicted brain age [cPRED] and corrected BAG [cBAG]), 9 ROIs survived multiple-comparisons analysis. These were the right middle frontal gyrus, right superior frontal gyrus medial segmesnt, right middle temporal gyrus, right putamen, left supplementary motor cortex, right central operculum, left cerebellum exterior, left central operculum, and the right cerebellum exterior (**Table 1**).

**Table 1.**
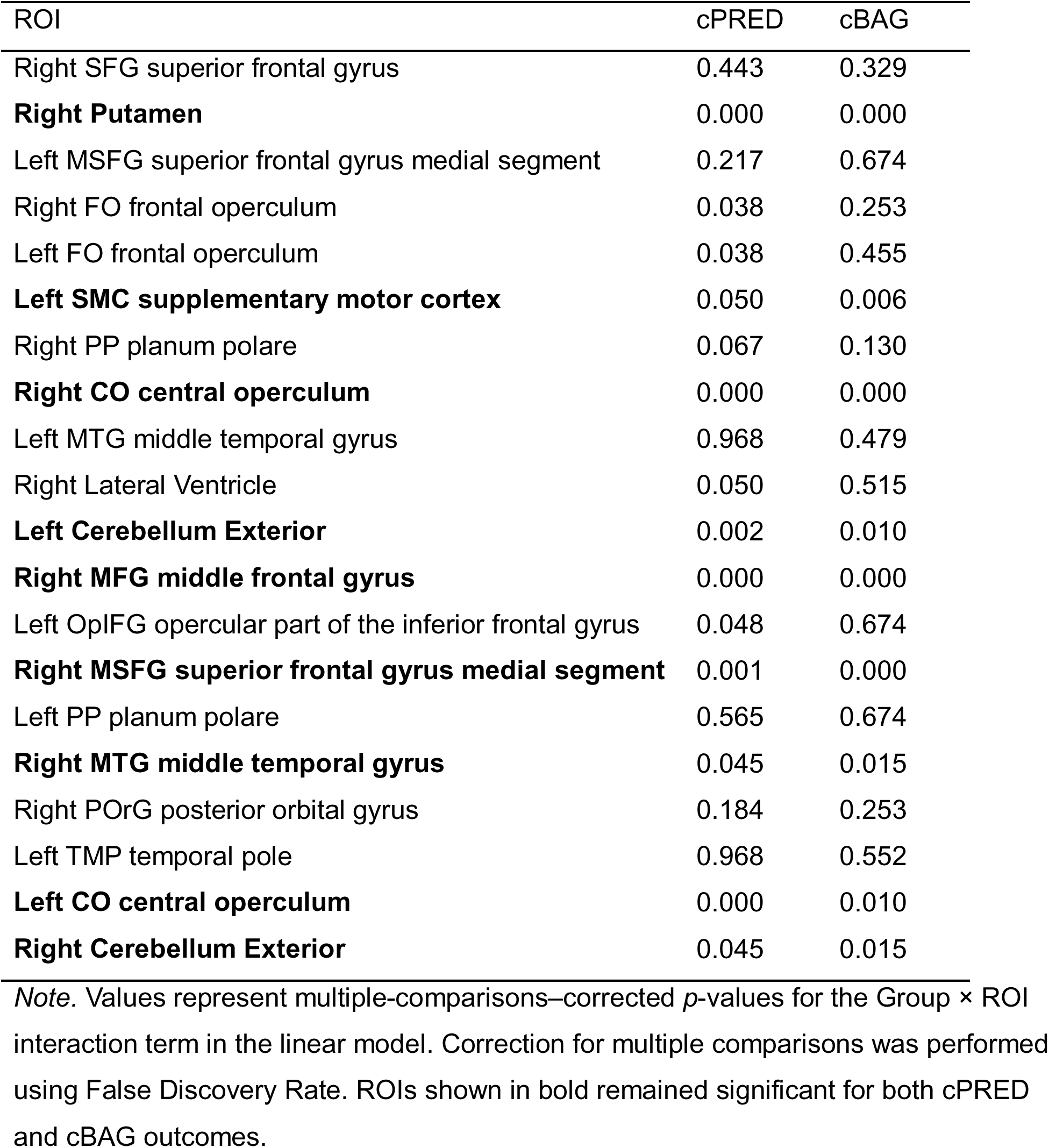
Multiple-comparison–corrected p-values for Group × ROI interaction effects on cPRED and cBAG.

### BAG analysis

The groupwise differences in outcome (BAG, cBAG, PRED, cPRED) are depicted in **Figures 1a** and **1b** with a general trend of divergence in early adulthood that increased into older adulthood across the various outcomes. Each plot includes a regression line per group to model the effect, with data collapsed across all sites.

**Figure 1:**
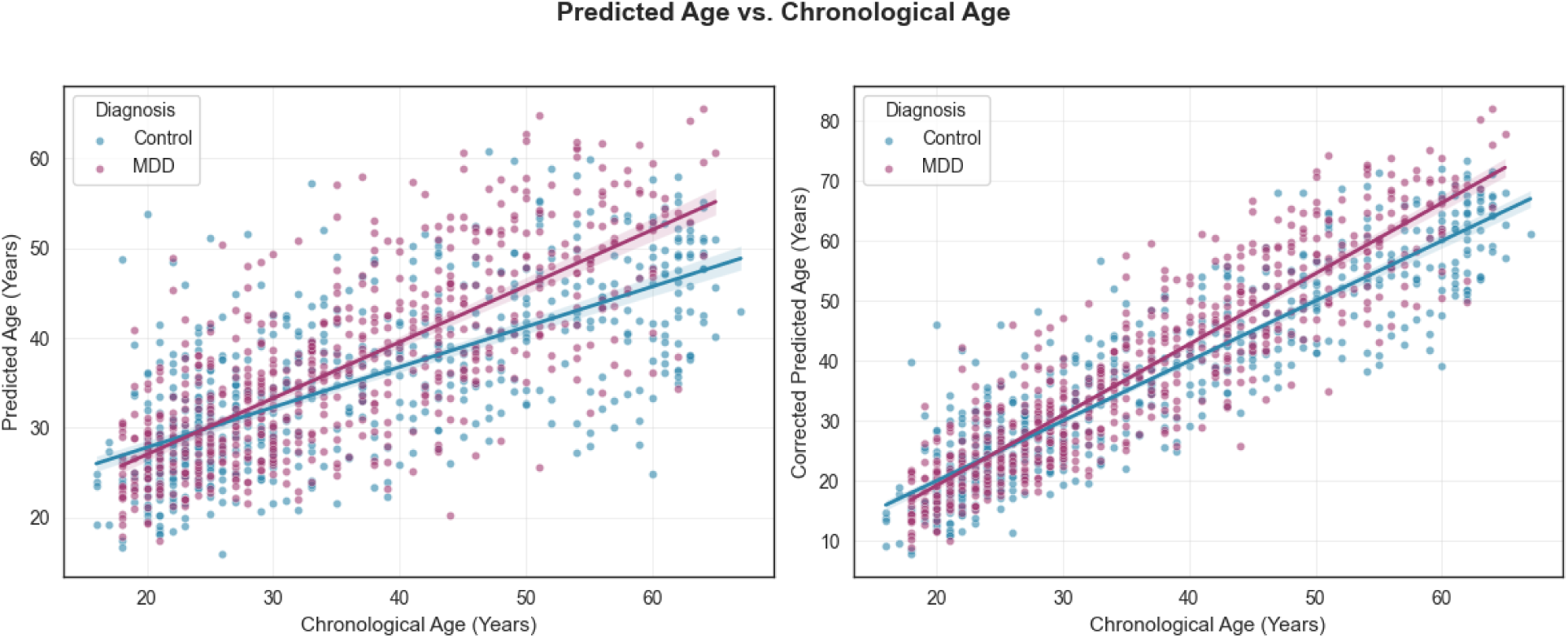
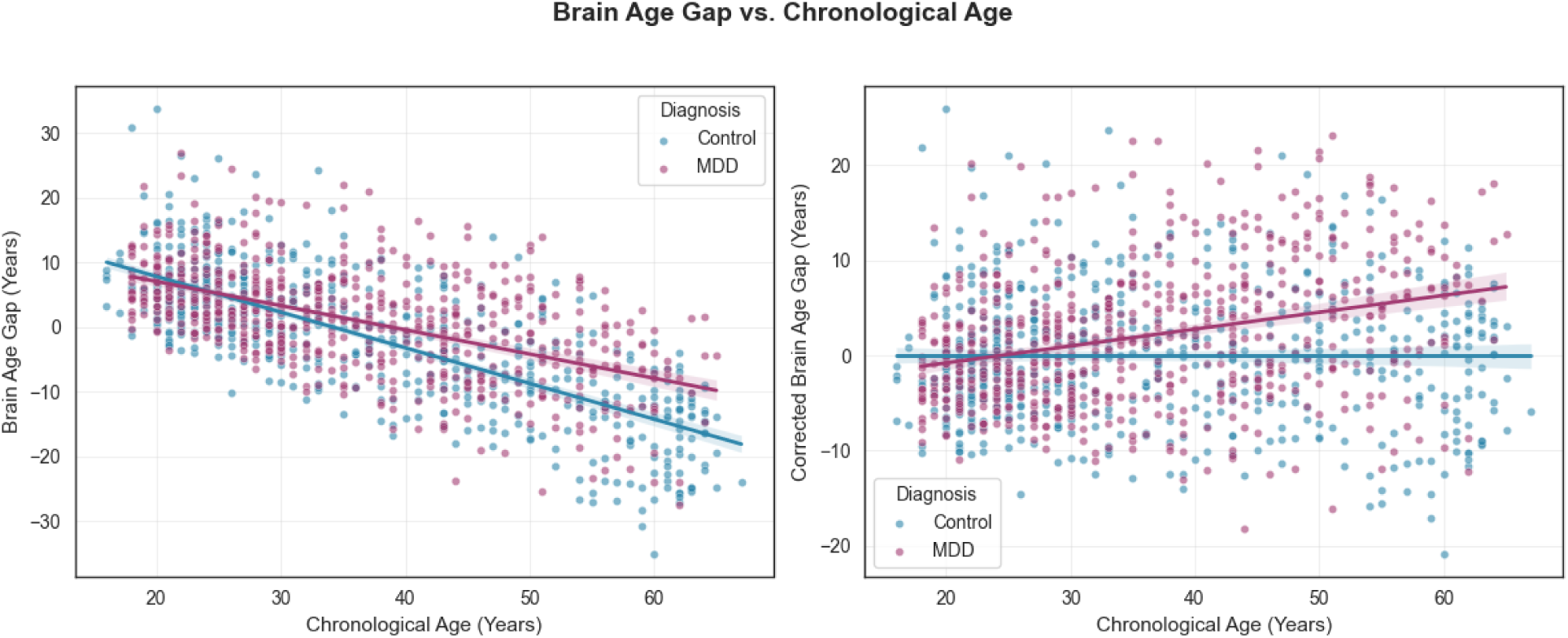
Plotting outcomes PRED and cPRED (Figure 1a) and BAG cBAG (Figure 1b) vs. chronological age, with regression lines (and associated 95% confidence intervals) for the MDD group (n=645) and control group (n=645) to demonstrate overall trends. Data across all sites are included in the above plots. Associations between age and brain age–related outcomes in individuals with major depressive disorder (MDD) and healthy controls. Scatterplots show (top left) predicted brain age (PRED), (top right) corrected predicted brain age (cPRED), (bottom left) brain age gap (BAG), and (bottom right) corrected brain age gap (cBAG) plotted against chronological age. Separate regression lines with 95% confidence intervals are shown for the MDD group (n = 645) and healthy control group (n = 645). Data from all contributing sites are included.

Across the entire cohort, MDD subjects showed a significantly elevated cBAG compared to controls (mean difference = 2.014 years, standard deviation 6.97 years; t = 5.591, p < 0.001, Cohen’s d = -0.26). Age-stratified analyses revealed that the group differences were not uniform across age ranges (**Figure 2**). While younger age groups (18-25 and 26-35 years) showed non-significant differences (p > 0.05), significant differences emerged in older age groups. Specifically, the 36–45-year age group showed a 2.88 year cBAG (p = 0.003, Cohen’s d = -0.39), whereas those aged 46-55 years showed a 3.63 year difference (p = 0.002, Cohen’s d = -0.45), and the 55+ year age group showed the cBAG of 6.85 years (p < 0.001, Cohen’s d = -0.94) (**Table 2**).

**Figure 2.**
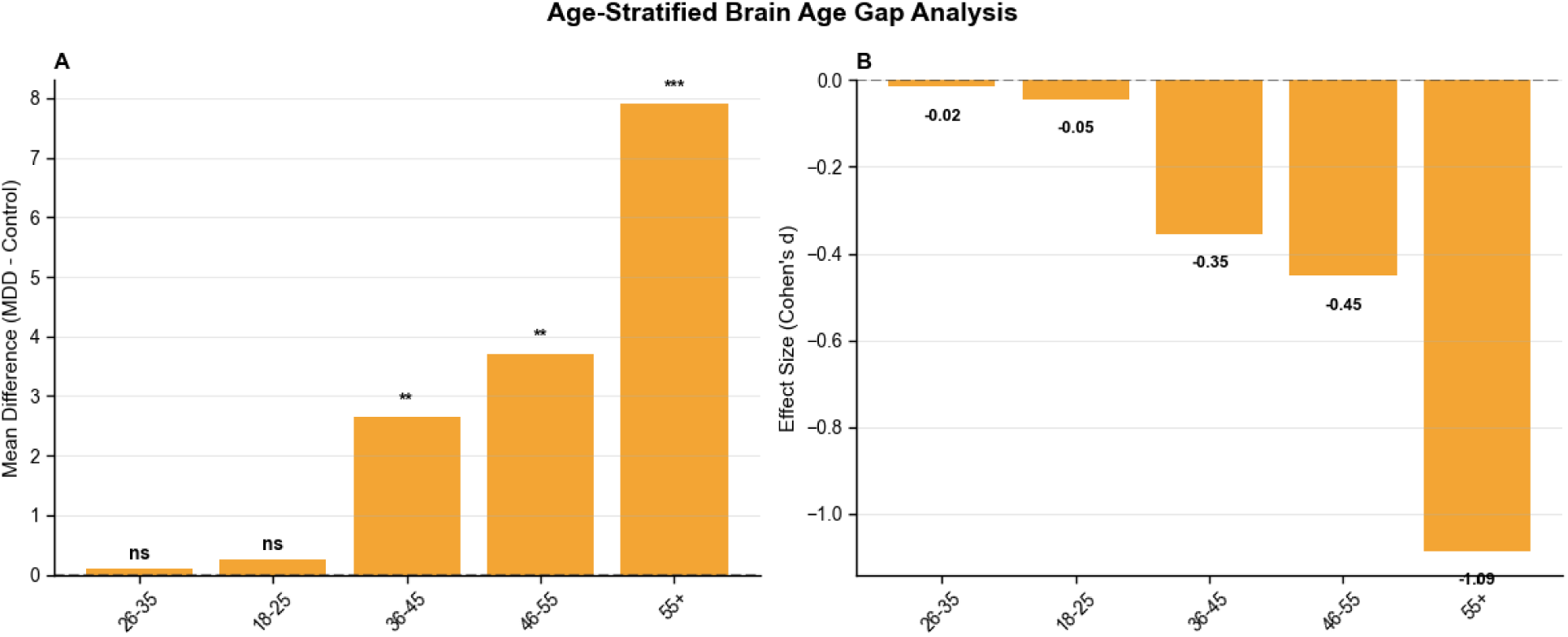
Age-stratified analysis of corrected brain age gap (cBAG). (A) Mean group difference in corrected brain age gap (MDD − control) across age bands. Bars represent mean differences, with significance indicated above each age group (ns, not significant; **p < .01; ***p < .001).(B) Corresponding effect sizes (Cohen’s *d*) for group differences in each age band. Negative values indicate higher cBAG in individuals with MDD relative to controls, based on group coding.

**Table 2.**
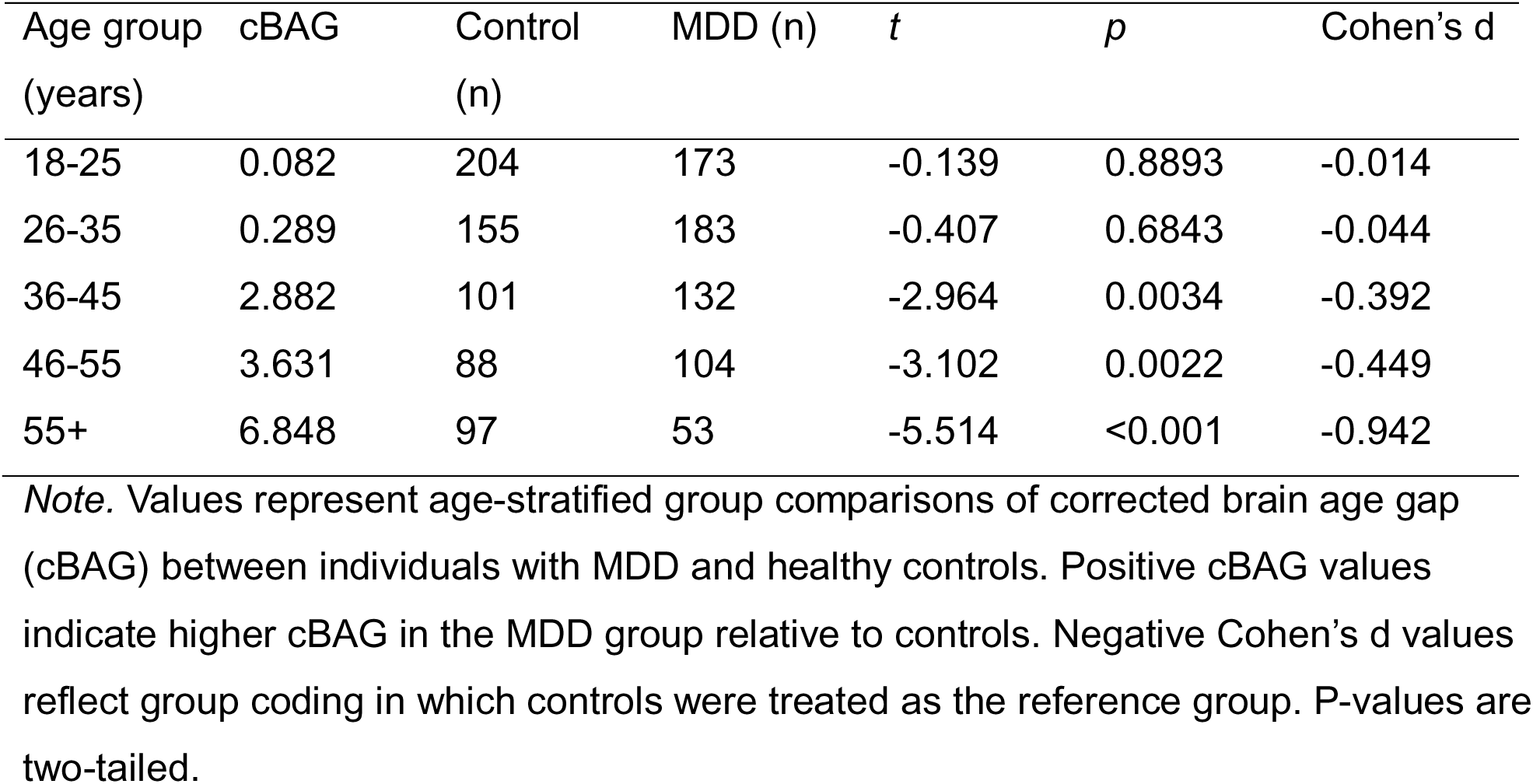
Age-stratified group differences in corrected brain age gap (cBAG)

**Table 3.** Demographics for the MDD and healthy control participants.

|  | Healthy controls | MDD Participants |
| --- | --- | --- |
| Number | 645 | 645 |
| Sex (% male) | 39.8 | 36.7 |
| Age | 36.5 ± 14.2 | 35.7 ± 12.3 |
| Education | 15.0 ± 2.8 | 13.4 ± 4.1 |
| Ethnicity (n, %) |  |  |
| *White* | 376, 58.3% | 237, 36.7% |
| *Asian* | 166, 25.7% | 237, 36.7% |
| *Black* | 16, 2.5% | 15, 2.3% |
| *Hispanic/Latino* | 8, 1.2% | 6, 0.9% |
| *Native American* | 4, 0.6% | 11, 1.7% |
| *Pacific Islander* | 1, 0.2% | 2, 0.3% |
| *Mixed* | 8, 1.2% | 10, 1.6% |
| *Missing* | 66, 10.2% | 194, 30.1% |

Percentage of male participants is presented. Mean age in presented in years with standard deviation (SD). Mean years of education and SD are presented. Ethnicity (self-report) and handedness are presented. Missing information is either because data were not collected or were not shared. Ethnicity for STRADL, Oxford, Remedi and KCL was estimated to be around 90%, 90%, 90% and 95% White respectively. The groups did not differ on age ( $t=1.09$ , $p=0.278$ ), sex distribution ( $\chi^2=1.18$ , $p=0.276$ ), or ethnicity ( $\chi^2=12.06$ , $p=0.099$ ), but did differ with respect to education ( $t=6.70$ , $p=0.000$ ) and handedness ( $\chi^2=31.56$ , $p=0.000$ ).

Prior work by our group identified two neuroanatomical MDD phenotypes, namely Dimension 1 (wherein white and grey matter was relatively preserved vs. controls) and Dimension 2 (characterized by subtle, widespread volume losses in both grey and white matter)^19^. When looking only at subjects belonging to these Dimensions, we found that the cBAG was 1.096 ± 7.201 years for those belonging to Dimension 1 and 2.779 ± 7.100 years for those in Dimension 2 (**Figure 3**). The difference in cBAG between these two sub-groups (1.634 years) was significant (t = -2.421, p = 0.015). Similarly, we found significant cBAG differences between Dimension 1 vs. healthy controls (t = -2.169, p = 0.03) and Dimension 2 vs. healthy controls (t = -6.031, p = < 0.001).

**Figure 3.**
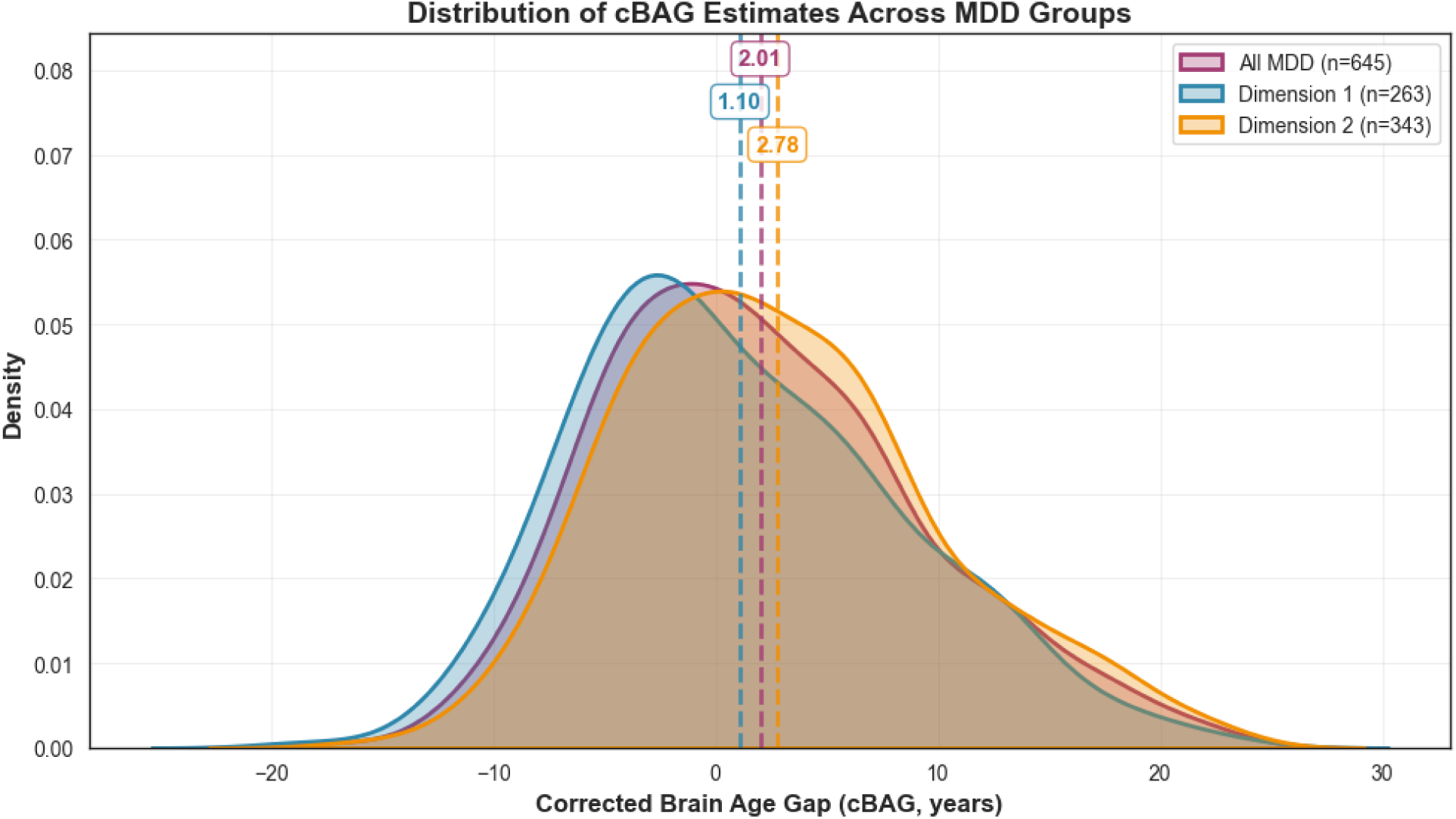
Corrected Brain Age Gap (cBAG) distributions across all MDD participants and the two MDD dimensions, representing distinct neuroanatomical MDD phenotypes. Distribution of corrected brain age gap across MDD dimensions. Density plots showing the distribution of corrected brain age gap (cBAG) for all individuals with major depressive disorder (MDD; n = 645) and for the two MRI-derived MDD dimensions (Dimension 1, n = 263; Dimension 2, n = 343). Dashed vertical lines indicate the mean cBAG for each group.

### Site effects

Site-specific analysis revealed heterogeneity across sites, with an intraclass correlation coefficient (ICC) of 0.111, indicating that 11.1% of the variance in brain age gap was attributable to site differences (F-statistic = 6.75, p < 0.001). The effect size for site as a fixed-effect was moderate (η^2^ = 0.055), explaining 5.5% of the total variance. Leave-one-site-out cross-validation (LOSO-CV) was performed to evaluate model generalizability, where the model was trained on data from 10 sites and tested on the remaining held-out site. This approach revealed variability in model performance across sites, with some sites showing higher MAE than others, and that one site (STRADL) generalizes less-well than others, per the LOSO-CV analysis (**Figures 4, 5**).

**Figure 4.**
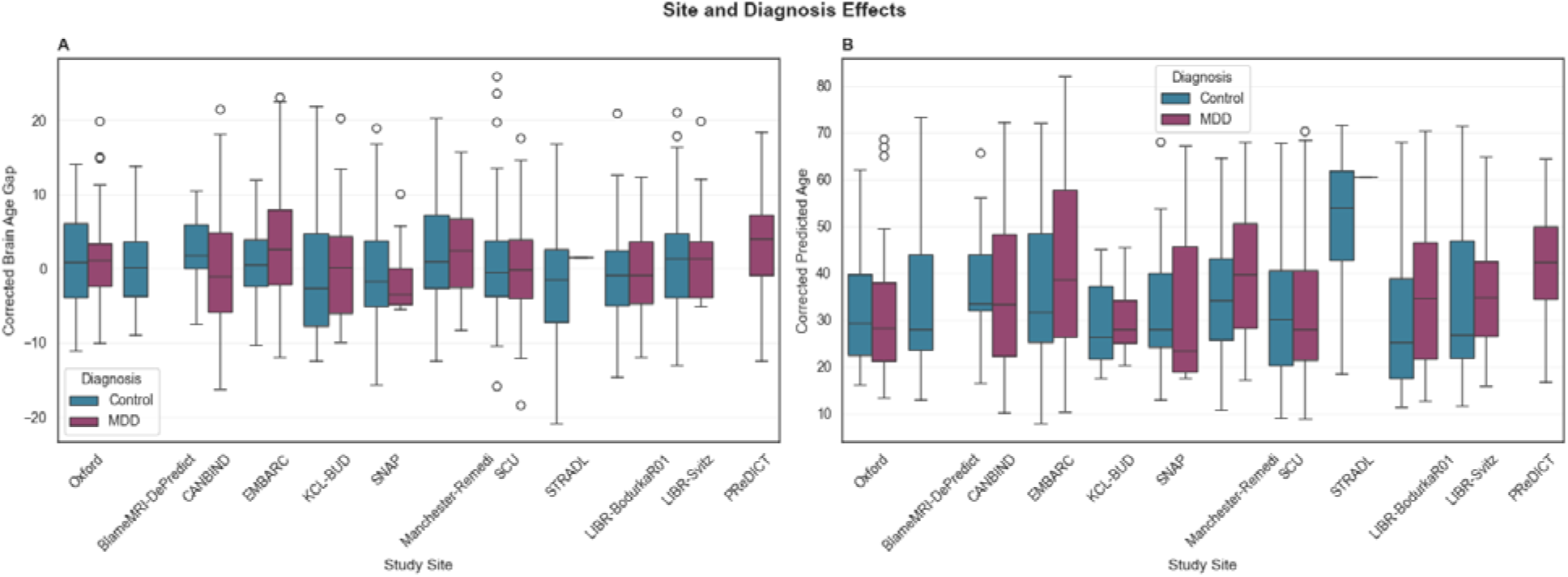
Site and diagnosis effects on brain age metrics for (A) cBAG and (B) cPRED by study site and diagnosis group. Boxplots show median (center line), interquartile range (box), and whiskers extending to 1.5× IQR. Each site is stratified by diagnosis: Control (blue) and Major Depressive Disorder (MDD, pink). The plots reveal both site-specific variation and diagnosis-related differences in brain age metrics across the multi-site sample.

**Figure 5.**
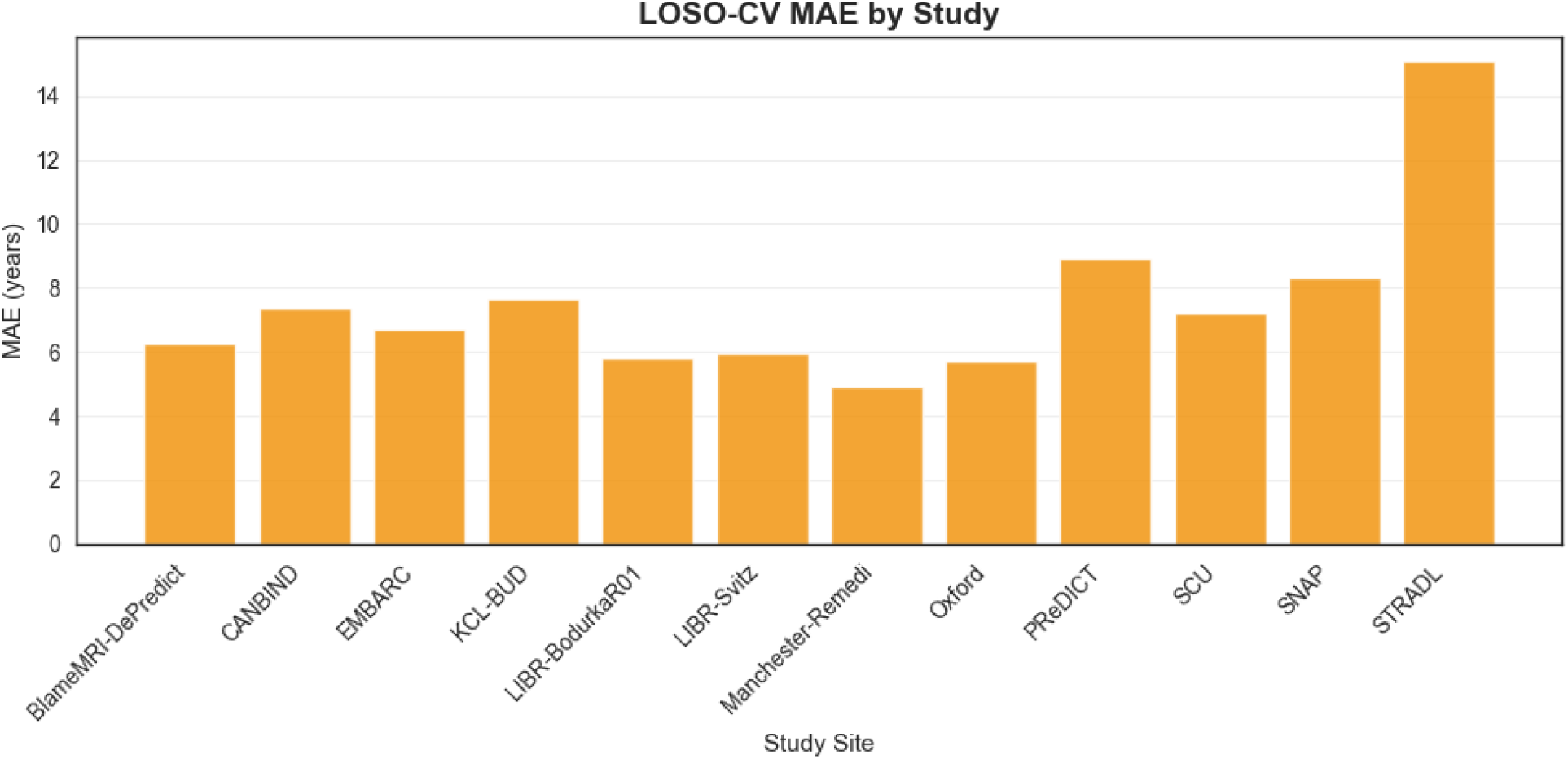
MAE in years when each site is left out of training. Bars show MAE on the held-out site; lower values indicate better generalization. This evaluates site-specific performance and model portability across sites.

## DISCUSSION

In the present large, harmonized, multi-site study, we found that MDD is associated with accelerated structural brain aging, reflected in an approximately 2-year increase in cBAG relative to healthy controls. Importantly, this effect was not uniform across the adult lifespan. No significant differences were observed in younger adults (18-25 and 26-35 years), but progressively larger increases emerged midlife and older age groups, with the largest difference reaching 6.85 years in MDD who were 55+ years. The findings demonstrate that accelerated brain aging in MDD is age contingent rather than age invariant, emerging after the mid-30s and becoming increasingly pronounced with advancing age. This pattern is consistent with neuroprogression models^27^, which propose that the biological effects of depression may accumulate over time and become more evident in later in the course of illness with advancing age.

Our results suggest a larger BAG than the most recent meta-analysis on the topic, which reported a mean difference of 0.95 (95% CI: 0.55, 1.35) and 1.12 (95% CI: 0.41, 1.83) per fixed- and random-effects analyses, respectively^10^. In contrast, the cBAG in our sample (inclusive of all age categories) was 2.01 years. Early work using support vector regression in a cross diagnostic cohort (n_Control_ = 800, n_MDD_ = MDD) reported a mean BAG of +4.0 (± 6.2) years^28^, whereas pilot studies (n_control_ = 40; n_MDD_ = 38) showed no significant gap^29^. Kauffman et al.^30^ used a model similar to ours (wherein they used xgboost and we used GradientBoostingRegression), and found a BAG of 0.86 (± 6.53) years in a sample of 208 healthy controls and an equivalent number of MDD cases. Larger consortia studies using ridge regression models trained on more than 3,000 healthy participants reported mean gaps of 0.99 years in the ENIGMA MDD sample (n=2126)^16^ and +2.8 years in the Netherlands Study of Depression and Anxiety cohort (n=285)^31^. Interestingly, large-scale fMRI studies provide converging results. A multisite fMRI analysis of 1,276 Chinese subjects reported a +4.4-year gap with stronger effects in antidepressant users^32^, with earlier fMRI brain age (n_MDD_ = 109) studies reporting a smaller gap of 2.11 years^15^. Within this context, our harmonised multisite study of 645 MDD cases and 645 matched controls used gradient boosting regression and nested cross-validation to identify a mean BAG of 2.01 years overall, though the differences were negligible in early adulthood yet increased to nearly 7-years in those aged over 55 years. These age-specific findings align with those reported in a brain age study by Christman et al.^13^ who stratified their cohort into mid-life (n=170, mean chronological age = 36.21 years) and older-adults (n=154, mean chronological age = 66.41 years), and found that the BAG was significantly greater in the older cohort. One of the main differences between our study and previous ones was the relatively small number of individuals with MDD above 65 years of age, which may account for the relatively larger cBAG found in this study, since our as well as previous studies have shown that BAG seems to widen with age.

Prior neuroimaging work from our group identified two neuroanatomical phenotypes, or Dimensions, of MDD^19^. This earlier work shows that subjects belonging Dimension 1 (n=290) were characterized by largely preserved gray and white matter, whereas those in Dimension 2 (n=395) demonstrated widespread volumetric reductions in both grey and white matter. Importantly, class membership predicted treatment response, with those in Dimension 1 experiencing significant symptom improvement after receiving SSRIs but not placebo^18^, as well as less cognitive impairment, adverse life events, self-harm and suicide attempts, and a more benign atherogenic metabolic profile^33^. Our present analysis shows that Dimension 1 subjects had a cBAG of 1.096 ± 7.201 years vs. 2.779 ± 7.100 for those in Dimension 2. The convergence of findings across these two independent analytical approaches, namely neuroanatomical clustering and brain age estimation, is consistent with a recent review of neuroimaging studies showing that parcelling the depression heterogeneity in imaging-based disease biotypes can improve the prediction of antidepressant treatment response toward personalized medicine^34^. The finding that Dimension 2, which has been associated with more severe neuroanatomical alterations and differential treatment response^19^, also exhibits the most pronounced brain age acceleration, raises the possibility that accelerated brain aging may be related to core pathophysiological mechanisms underlying this particular MDD biotype. Future longitudinal studies should examine whether brain age acceleration predicts treatment response or clinical course within these dimensional subtypes.

With respect to individual ROIs, several survived multiple comparisons testing (Table 1). The right putamen contributed the most to the BAG prediction (per the absolute SHAP value), a region also found to contribute to brain age prediction in MDD subjects with anhedonia^35^. Notably, prior reports have noted that the putamen volume declines nearly twice as fast with age in MDD in comparison to controls^36^, and that reduced putamen volumes are associated with high-risk of developing depression^37^. The feature with the second highest impact on the BAG prediction was the left supplementary motor cortex, which has been associated with a reduced cortical thickness in MDD per a recent meta-analysis^38^. The right central operculum was the third most impactful feature on BAG. A separate meta-analysis of voxel-based morphometry neuroimaging studies showed that this region was part of a broader cluster that differed between healthy controls and those with MDD^39^. Collectively, our results are consistent with prior reports on the neurobiology of depression, though they do not yet provide a mechanistic understanding for why these changes are observed. The field would benefit from future studies that incorporate multi-modal imaging for concurrent brain structure and function assessment within the same sample. As well, brain age studies are encouraged to report ROIs that drive BAG, as this may provide additional insights into which regions are contributing to neurophysiological differences in MDD across different cohorts. Beyond individual regions, at a circuit or network level, the ROIs that significantly drive the BAG are implicated with striatal, frontal, opercular, temporal, and cerebellar functioning, which include cognitive control, reward processing, sensorimotor integration, and affect regulation^40–43^.

From a methodological perspective, compared with prior research, our approach offers several strengths. First, we examined a highly phenotyped sample of non-treatment resistant, medication-free subjects with MDD, minimizing current pharmacological confounds on brain age estimations. Many earlier large-scale efforts relied on aggregated summary statistics from individual sites, which limited harmonization and reduced the ability to explore site effects. In contrast, we obtained deidentified, individual-level structural MRI data from 11 cohorts and applied a fully automated MUSE segmentation pipeline to extract 145 ICV-normalized ROI volumes for each participant^21^. This individual-level approach eliminates reliance on summary statistics and allows for harmonization of multi-site data using methods such as ComBat-GAM^21^. This is important as ComBat-based harmonization approaches have been shown to reduce the impact of unwanted site effects while preserving biological signal^44^. Site effects were explicitly modelled, with leave-one-site out validation showing that site accounted for only ∼11 % of variance. Further, we applied age-bias correction and propensity-score matching to ensure that controls and MDD subjects were balanced on age, ICV and sex. Our analytic pipeline employed nested cross-validation with a gradient boosting regressor (rather than linear ridge regression, as used in prior large-scale studies) to optimize hyperparameters and avoid overfitting. Further, to our knowledge, our study is the first to use SHAP values to understand the contribution of individual ROIs to the brain age prediction, which adds explainability and insight into which neuroanatomical regions are driving differences are driving the BAG in MDD. This allowed us to uncover significant Group X ROI interactions that were not captured in prior summary-level studies. Altogether, we feel our work continues to build on BAG research in MDD with a large sample and robust methodology.

Despite these strengths, there are limitations to this study that warrant consideration. First, despite rigorous harmonization and propensity matching, residual site effects and sample heterogeneity cannot be fully excluded. Second, the cross-sectional design limits inference on causality and trajectories of brain age acceleration. Finally, while GradientBoostingRegression provides robust predictions, alternative machine learning approaches and multimodal imaging data may capture additional aspects of accelerated aging in MDD that our study did not.

## CONCLUSIONS

This large, harmonized, multi-site study provides evidence that MDD is associated with accelerated brain aging, with effects that grow progressively stronger after mid-30s. This study also found that BAG is most pronounced in a neuroanatomical phenotype associated with poorer antidepressant treatment response, cognitive impairment and worse key clinical outcomes. Regional contributions from striatal, frontal, opercular, temporal, and cerebellar circuits suggest that MDD disproportionately affects networks supporting reward, cognitive control, and affect regulation. These findings align with neuroprogression models, which posit that MDD involves cumulative biological changes over time, and emphasize the potential utility of the BAG as a biomarker for disease burden and outcome.

**Supplementary figure 1.**
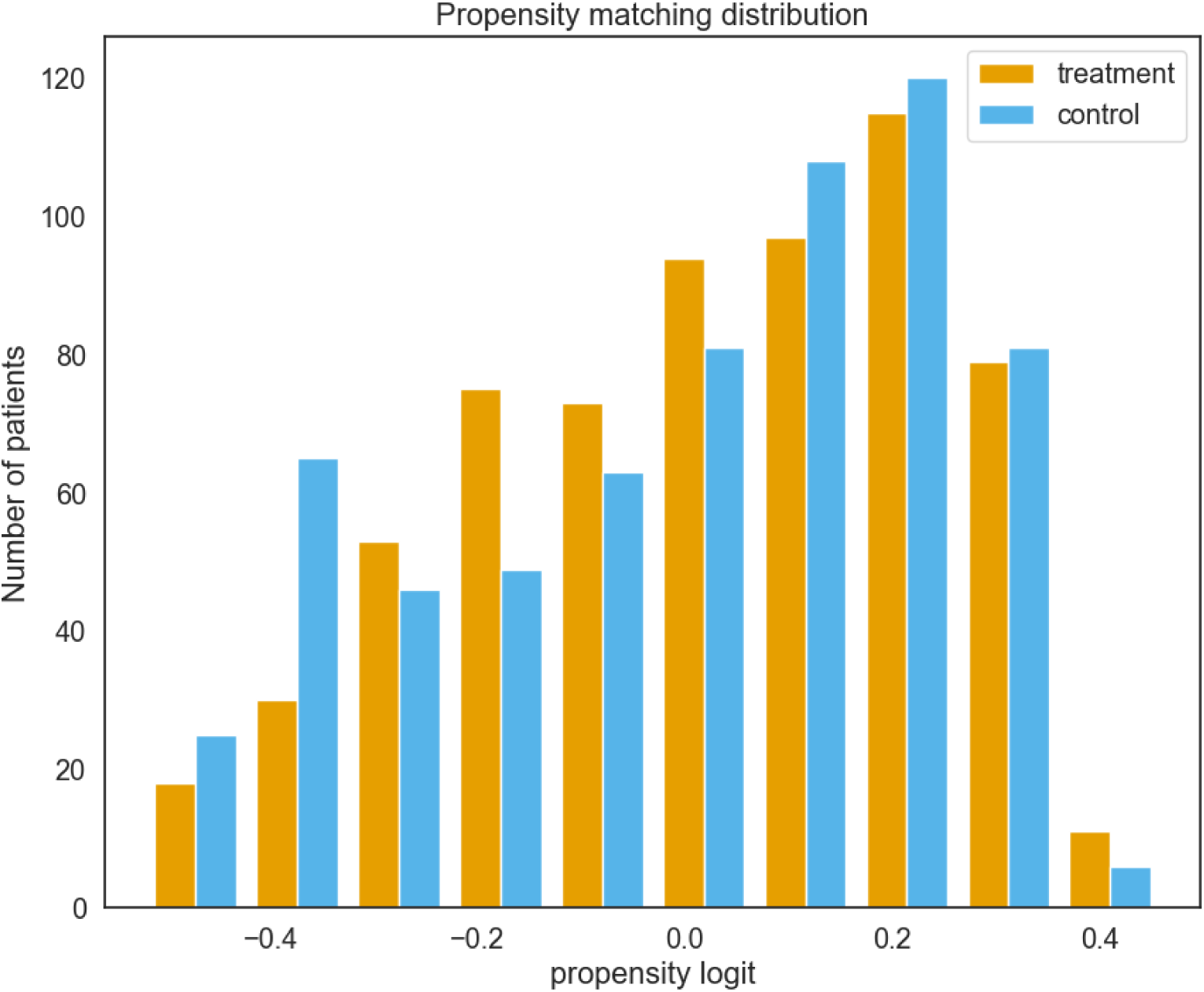
Distribution of propensity scores across treatment and control groups following matching.

**Supplementary table 1.** Mean absolute error and hyperparameters of the brain age prediction model across outer cross-validation folds.

| Parameter | Outer fold 1 | Outer fold 2 | Outer fold 3 | Outer fold 4 | Outer fold 5 |
| --- | --- | --- | --- | --- | --- |
| MAE (years) | 8.129 | 7.519 | 6.788 | 7.066 | 7.391 |
| Learning rate | 0.113 | 0.086 | 0.086 | 0.063 | 0.036 |
| Loss function | Squared error | Absolute error | Absolute error | Squared error | Squared error |
| Max tree depth | 4 | 3 | 3 | 4 | 2 |
| Estimators (n) | 221 | 268 | 268 | 468 | 289 |
| Minimum samples split | 8 | 14 | 14 | 5 | 13 |
| Minimum samples leaf | 5 | 3 | 3 | 5 | 5 |
| Subsample | 0.662 | 0.670 | 0.670 | 0.903 | 0.760 |

**Supplementary Table 2.** Sample distribution by site and diagnostic group in the propensity-matched cohort.

| Site / Cohort | Control (n) | MDD (n) |
| --- | --- | --- |
| CAN-BIND | 17 | 76 |
| EMBARC | 37 | 233 |
| KCL | 20 | 20 |
| LIBR | 141 | 54 |
| Manchester-BLAME | 46 | 0 |
| Manchester-REMEDI | 30 | 40 |
| Oxford Cohort | 31 | 39 |
| PReDICT | 0 | 63 |
| SCU | 139 | 111 |
| SNAP | 50 | 8 |
| STRADL | 134 | 1 |

## Data Availability

All data produced in the present study are available upon reasonable request to the authors.

